# Aortic Valve Level as a Candidate Zone 4/5 Boundary Landmark in Endovascular Aortic Repair

**DOI:** 10.64898/2026.07.20.26358310

**Authors:** Tatsuya Nishii, Hiroki Horinouchi, Akiyuki Kotoku, Shumpei Mori, Yasutoshi Ohta, Tetsuya Fukuda

**Affiliations:** Department of Radiology, National Cerebral and Cardiovascular Center, Suita, Osaka, Japan; Division of Cardiology, Department of Medicine, David Geffen School of Medicine at University of California Los Angeles (UCLA), Los Angeles, California, USA

**Keywords:** Computed tomography angiography, Aortic Diseases, Aortic Aneurysm, Aortic Dissection, Endovascular Aneurysm Repair, Spinal Cord Ischemia, Adamkiewicz artery

## Abstract

**Background:** The zone 4/5 boundary has been described as the mid-descending aorta–T6 level, but this correspondence has uncertain anatomic support.

**Purpose:** To determine whether the aortic valve (AV) level approximates the descending aortic midpoint and to compare candidate boundaries relative to the critical segmental artery (CSA) origin.

**Materials and Methods:** This retrospective study included 204 patients who underwent Adamkiewicz artery-specific CT angiography from January 2022 through February 2026. The CSA was defined as the aortic origin of the segmental artery directly or collaterally connected to the Adamkiewicz artery. Along the descending-aortic centerline, the descending aortic midpoint was the point halfway between a site 20 mm distal to the left subclavian artery origin and the celiac artery origin; the AV-level midpoint was the point halfway between the centerline intersections of the left coronary and noncoronary aortic sinus planes. Equivalence between midpoints was tested within a prespecified ±20-mm margin. Distal CSA classifications were compared using McNemar tests.

**Results:** The study included 204 patients (median age, 72 years [IQR, 59–79 years]; 142 men and 62 women). The mean difference between the AV-level midpoint and descending aortic midpoint was -1.2 mm (90% CI, -3.3 to 0.9 mm; P < .001 for equivalence). The T6 vertebral level was 59.0 mm proximal to the descending aortic midpoint. The CSA origin was distal to the AV-level midpoint and descending aortic midpoint in 96.6% (197/204) and 95.6% (195/204), respectively (P = .68), but distal to the noncoronary aortic sinus plane in 89.7% (183/204; P = .001 versus the AV-level midpoint).

**Conclusion:** The AV-level midpoint approximated the descending aortic midpoint, whereas the T6 vertebral level was more proximal. Fewer critical segmental artery origins were classified as distal to the noncoronary aortic sinus plane than to the AV-level midpoint.

## Introduction

The aortic zone framework, commonly referred to as Ishimaru zones(1), has become standardized guideline terminology(2–4) for thoracic endovascular aortic repair (TEVAR). Although first applied mainly to proximal landing zones, this framework is now increasingly used to describe distal aortic coverage, where extensive descending thoracic coverage is associated with spinal cord ischemia (SCI)(5–7). The zone 4/5 boundary is therefore clinically important, but it remains less consistently defined than the proximal arch zones(8). Guidelines describe this boundary as “mid-descending aorta– T6”(2–4), without specifying whether the geometric midpoint coincides with T6 or providing a clear anatomic basis. This ambiguity is important when zone assignment depends on reproducible image-based definitions(9).

The origin of the Adamkiewicz artery (AKA) is an anatomically relevant reference for evaluating this boundary because the AKA commonly arises in the lower thoracic region(10) and contributes to the spinal collateral network(11). Although routine preoperative AKA identification remains debated, coverage of the segmental artery supplying the AKA may reduce spinal cord perfusion reserve in selected high-risk patients(12). Advances in noninvasive AKA imaging(10,13–15), including low-tube-potential acquisition(16) and high-spatial-resolution reconstruction(17), now allow AKA-specific CT angiography (AKA-CTA) to identify the AKA in 90%–97%(16–18) of candidates for aortic repair. These advances make the AKA origin a feasible anatomic reference for evaluating candidate definitions of the zone 4/5 boundary.

Neither conventional landmark is readily usable during TEVAR. The descending aortic midpoint requires CT centerline analysis and is not directly visible fluoroscopically, whereas the T6 vertebral level is a projected skeletal reference affected by working/viewing angle and vertebral counting. This procedural gap led us to evaluate the aortic valve (AV) level as an alternative landmark. Because a catheter or guidewire is often placed in the aortic root, the AV level can be localized fluoroscopically. This study aimed to determine whether the AV-level midpoint, defined as the centerline midpoint between the left coronary and noncoronary aortic sinus planes, approximates the descending aortic midpoint used as the conventional zone 4/5 boundary. We also assessed the relationship between the T6 vertebral level and the mid-descending boundary and compared candidate boundaries with the AKA origin on AKA-CTA.

## Materials and Methods

### Study Design and Patients

This retrospective single-center study reviewed consecutive patients who underwent AKA-CTA before aortic repair from January 2022 through February 2026. The institutional review board approved the study (approval No. R19039-4) and waived written informed consent because of its retrospective design. Of 204 patients, 102 were included in a previous study of the added value of AKA-CTA for TEVAR planning(18); all landmarks were newly measured for the present analysis, which had a different primary endpoint.

Eligible patients had an identifiable critical segmental artery (CSA) origin, defined below. Exclusion criteria were nonidentifiable AKA or predominant spinal cord perfusion through collateral pathways originating from the subclavian or iliac arteries.

Study size was justified on the basis of estimated precision for the primary endpoint. Preliminary data from 162 patients showed a 17.7-mm standard deviation in the AV-level versus descending aortic midpoint difference. To achieve a 95% confidence interval (CI) half-width of 2.5 mm, the cohort was expanded to at least 195 patients.

### Image Acquisition

AKA-CTA was acquired on a 192-row dual-source CT scanner (SOMATOM Force; Siemens Healthineers) in dual-power mode with automatic low-kV selection (70–100 kV). Non-contrast, early arterial, and late arterial scans covered the aortic arch to L4. Contrast material was delivered at a weight-adjusted iodine delivery rate of 26 mg I/kg/s, and timing was optimized in the descending aorta using the timing-bolus method. Images were reconstructed with a 160-mm field of view and a sharp filter kernel (Bv59); bone-subtraction images were generated(17). Detailed parameters are provided in Table S1.

### Image Analysis

Image analysis was performed on a workstation (Ziostation2, Ziosoft). Two board-certified radiologists (T.N. and A.K.) with 17 and 12 years of experience, respectively, independently identified the AKA and CSA with the previously reported stepwise 6-point scoring system(16,17). Consensus resolved discrepancies. The CSA origin was defined as the aortic origin of a segmental artery that directly gave rise to the AKA or, when that artery was occluded, of a segmental artery connected to the AKA through collateral pathways(17,19). The largest candidate CSA was selected when multiple candidate CSAs were present.

One radiologist (T.N.) measured distances on a semiautomatically generated, manually corrected arch-to-abdominal aortic centerline; for patients with dissection, the centerline followed the true lumen. Landmarks were branch origins (left subclavian artery, CSA, and celiac artery) and axial planes (mid-T6, mid-T9, left coronary aortic sinus, and noncoronary aortic sinus). Sinus planes were axial levels of the upper margin of the left coronary aortic sinus and lower margin of the noncoronary aortic sinus, respectively. Branch landmarks were assigned on orthogonal reformations; axial-plane landmarks, were assigned at centerline intersections with corresponding z-axis coordinates. Details are provided in Appendix S1 and Figure S1.

The descending aorta, defined by guideline zone 4 and 5 limits(4), extended from 20 mm distal to the left subclavian artery origin to immediately proximal to the celiac artery origin. The descending aortic midpoint was the segment centerline midpoint; the AV-level midpoint was midway between the two aortic sinus-plane centerline intersections. Candidate zone 4/5 boundaries were the descending aortic midpoint, AV-level midpoint, and T6 plane; aortic sinus planes were secondary AV-level references. The exploratory T8 upper-border estimate is detailed in Appendix S1. CSA-to-landmark distance equaled CSA origin distance minus landmark distance; positive values indicated a distal CSA origin. Relative distances were normalized to 0, 1, and 2 at the zone 4 start, descending aortic midpoint, and celiac origin, respectively.

### Outcomes

The centerline-derived descending aortic midpoint, a continuous, reproducible form of the mid-descending definition, served as the reference boundary. The primary outcome was the paired centerline- distance difference between the AV-level midpoint and descending aortic midpoint, calculated as AV- level midpoint distance minus descending aortic midpoint distance. Secondary outcomes were the T6 plane-descending aortic midpoint difference, the distance from each boundary to the CSA origin, and whether the CSA origin was distal to each landmark. Exploratory outcomes evaluated whether the descending aortic midpoint, AV-level midpoint, and T9 plane reflected CSA origin-distance variation in the T9-origin subset, defined as patients with a right or left ninth-intercostal CSA origin.

### Measurement Reproducibility

Interobserver and intraobserver measurements were performed in the last 25 cases and focused on four descending aortic centerline landmarks: AV-level midpoint, descending aortic midpoint, CSA origin, and T6 plane. A second radiologist (H.H.; 14 years of experience) performed measurements for interobserver assessment. The first observer (T.N.) repeated measurements at least 3 months later for intraobserver assessment.

### Statistical Analysis

Continuous variables were summarized as means with standard deviations or medians with interquartile ranges (IQRs). Analyses used R (version 4.6.0, R Foundation for Statistical Computing); P < .05 indicated statistical significance.

Equivalence between the AV-level and descending aortic midpoints was assessed with two one-sided tests using a prespecified ±20-mm margin and was concluded when the 90% CI for the paired difference lay entirely within this margin. This margin reflected a practical TEVAR-planning tolerance because thoracic stent-graft lengths are discrete and reported distal landing-zone deployment error is approximately 10 mm (IQR, 6.5–16.0 mm)(20); therefore, a 20-mm difference was considered unlikely to materially alter device-length selection or landing-zone planning. Multivariable linear regression explored clinical factors associated with the AV-level versus descending aortic midpoint difference. Individual agreement was summarized with 95% limits of agreement and the proportion of absolute differences ≤ 20 mm.

Distal CSA classifications were compared using paired McNemar tests with Holm adjustment. In the T9-origin subset, separate linear regression models assessed associations between CSA origin distance and each candidate landmark distance: T9 vertebral-level, descending aortic midpoint, and AV- level midpoint. Bootstrap comparisons evaluated standardized beta differences. Distance-measurement reproducibility was evaluated using intraclass correlation coefficients.

## Results

Among 221 consecutive patients who underwent AKA-CTA before aortic repair, the AKA was identified in 95.5% (211/221), and 204 were included after exclusion of patients with predominant subclavian or iliac collateral supply (Figure 1). Median age was 72 years (IQR, 59–79 years), and 69.6% (142/204) were men. Underlying aortic disease was dissection in 62.3% (127/204) and aneurysm in 30.9% (63/204) (Table 1). The CSA arose from the aorta at or caudal to the eighth intercostal level in 198/204 patients (97.1%; exact 95% CI, 93.7% to 98.9%) (Figure 2).

**Figure 1.**
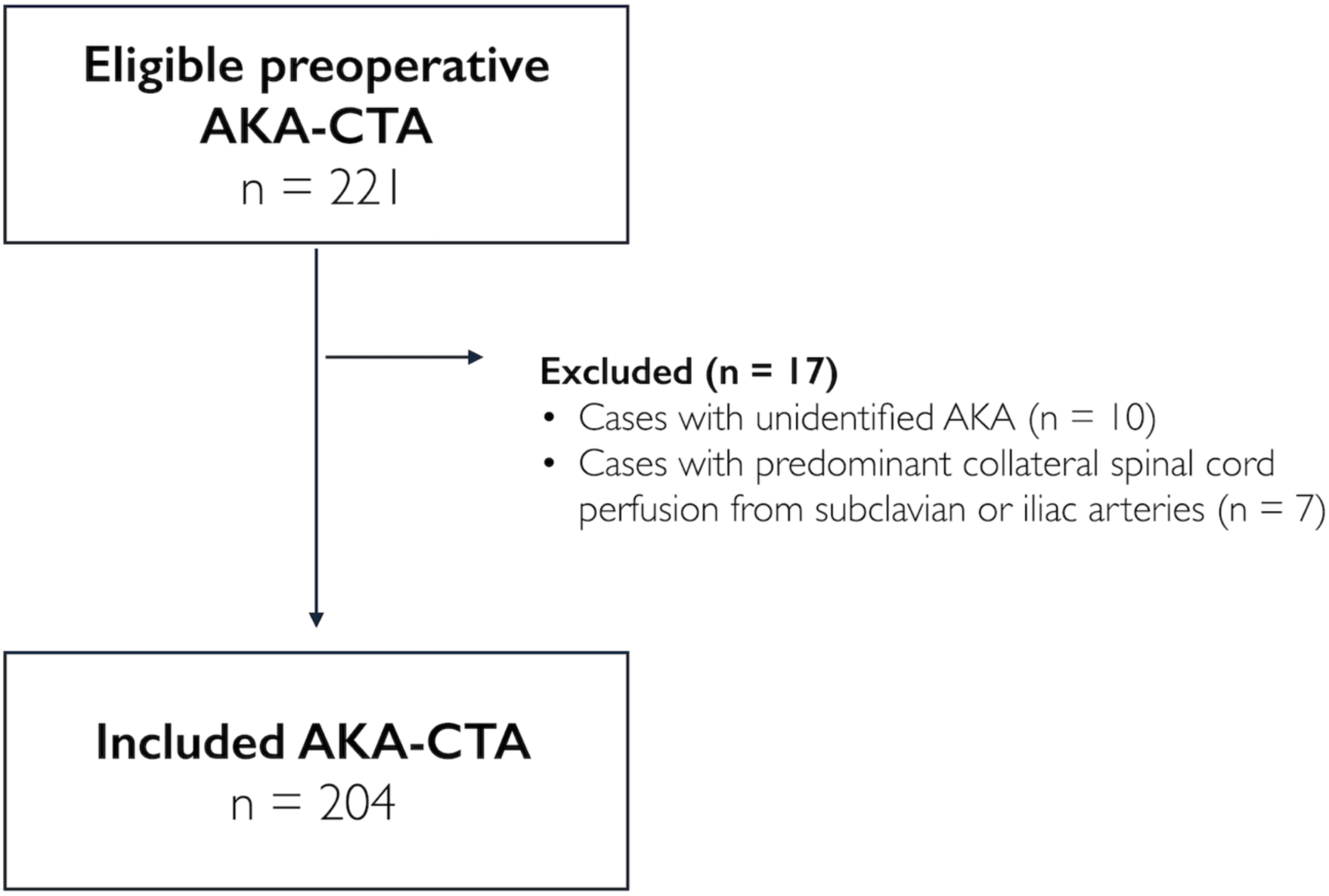
Flowchart of patients in this study. AKA = Adamkiewicz artery, AKA-CTA = AKA-specific CT angiography.

**Figure 2.**
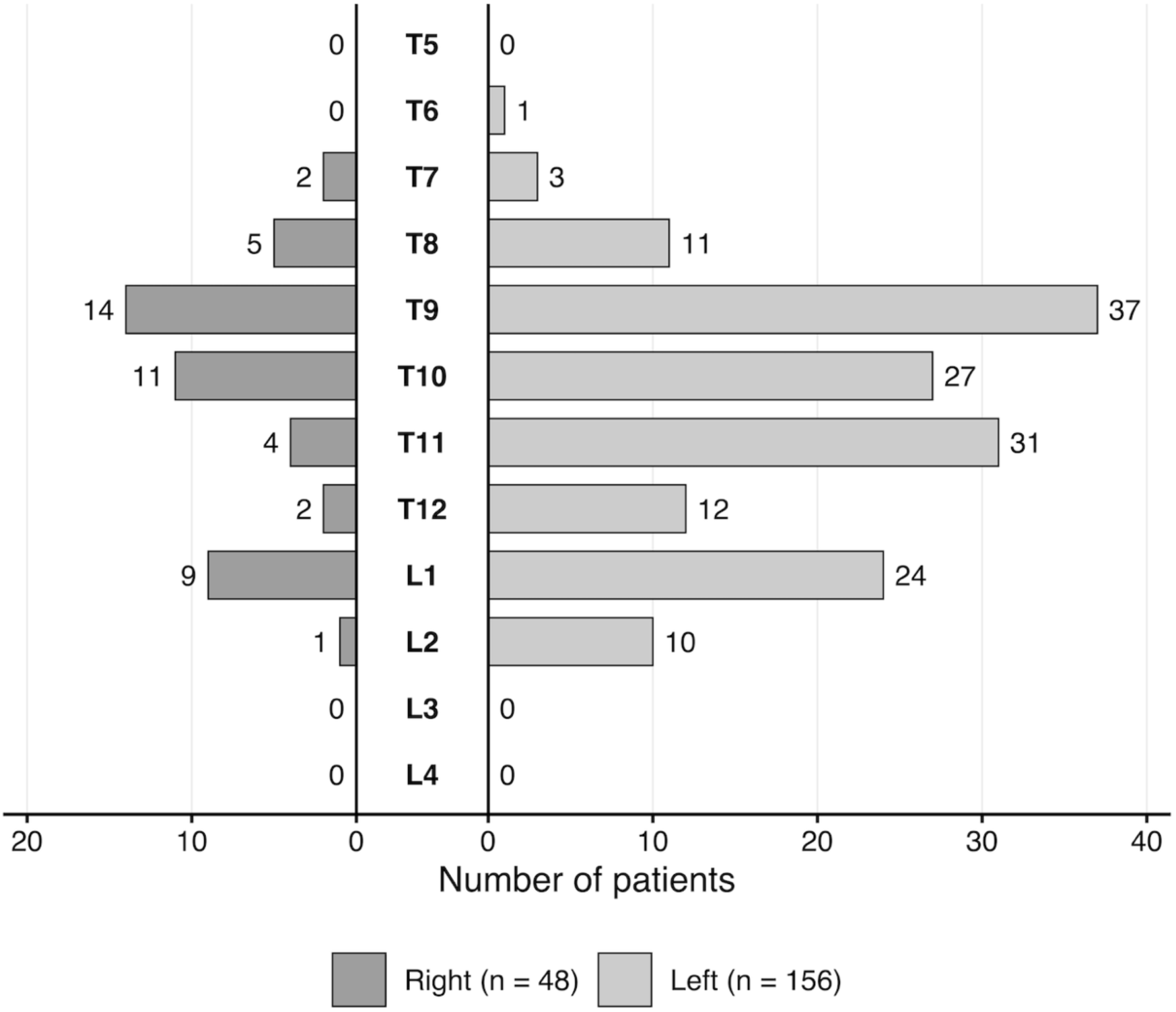
Distribution of CSA origins on Adamkiewicz artery-specific CT angiography. CSA = critical segmental artery.

**Table 1.** Patients’ characteristics and Adamkiewicz artery-specific CT angiography findings.

| Variable | N = 204 |
| --- | --- |
| Age * | 72 (59, 79) |
| Sex |  |
| Female | 62 (30.4) |
| Male | 142 (69.6) |
| Height (cm) * | 166 (158, 171) |
| Weight (kg) * | 62.0 (51.8, 72.1) |
| Body mass index (kg/m<sup>2</sup>) * | 22.7 (20.3, 25.3) |
| Disease |  |
| Dissection | 127 (62.3) |
| Aneurysm | 63 (30.9) |
| Intramural hematoma | 6 (2.9) |
| Other | 8 (3.9) |
| Previous proximal aortic repair | 136 (66.7) |
| *Scan parameters* |  |
| Contrast material |  |
| 370 mgI/mL | 167 (81.9) |
| 350 mgI/mL | 37 (18.1) |
| Injection rate (mL/sec) * | 4.3 (3.9, 4.6) |
| Contrast material volume (mL) * | 87 (79, 89) |
| Tube potential |  |
| 70 kV | 110 (53.9) |
| 80 kV | 58 (28.4) |
| 90 kV | 29 (14.2) |
| 100 kV | 7 (3.4) |
| CT dose index for arterial phase (mGy) * | 16.6 (14.2, 23.0) |
| Dose-length product for arterial phase (mGy×cm) * | 722.1 (595.1, 1021.8) |
| Scan length (mm) * | 425.8 (400.9, 450.6) |
| Total dose length product (mGy×cm) * | 1888.8 (1515.6, 2575.5) |
| *Adamkiewicz artery-specific CT angiography findings* |  |
| Hairpin curve presence | 204 (100) |
| Continuity score |  |
| Score 1 | 13 (6.4) |
| Score 2 | 47 (23.0) |
| Score 3 | 144 (70.6) |
| Washout positive | 187 (91.7) |
| Branching lumen |  |
| True lumen | 170 (83.3) |
| False lumen | 29 (14.2) |
| Both lumen | 5 (2.5) |
| Collateral pathway | 25 (12.3) |

Note.—Data represent the numbers of patients, with percentages in parentheses.
\* Data are presented as medians, with interquartile ranges in parentheses.

### Centerline Distances of Anatomical Landmarks

Landmark centerline distances are summarized in Table 2 and Figure 3. Mean distances to the AV-level and descending aortic midpoints were 131.1±29.5 mm and 132.2±18.2 mm, respectively; the T6 plane was more proximal. The CSA origin was at a median distance of 207.5 mm (IQR, 156.5–271.6 mm). Relative-distance plots showed AV-level midpoint clustering around the descending aortic midpoint and a more distal CSA origin distribution (Figure 3A).

**Figure 3.**
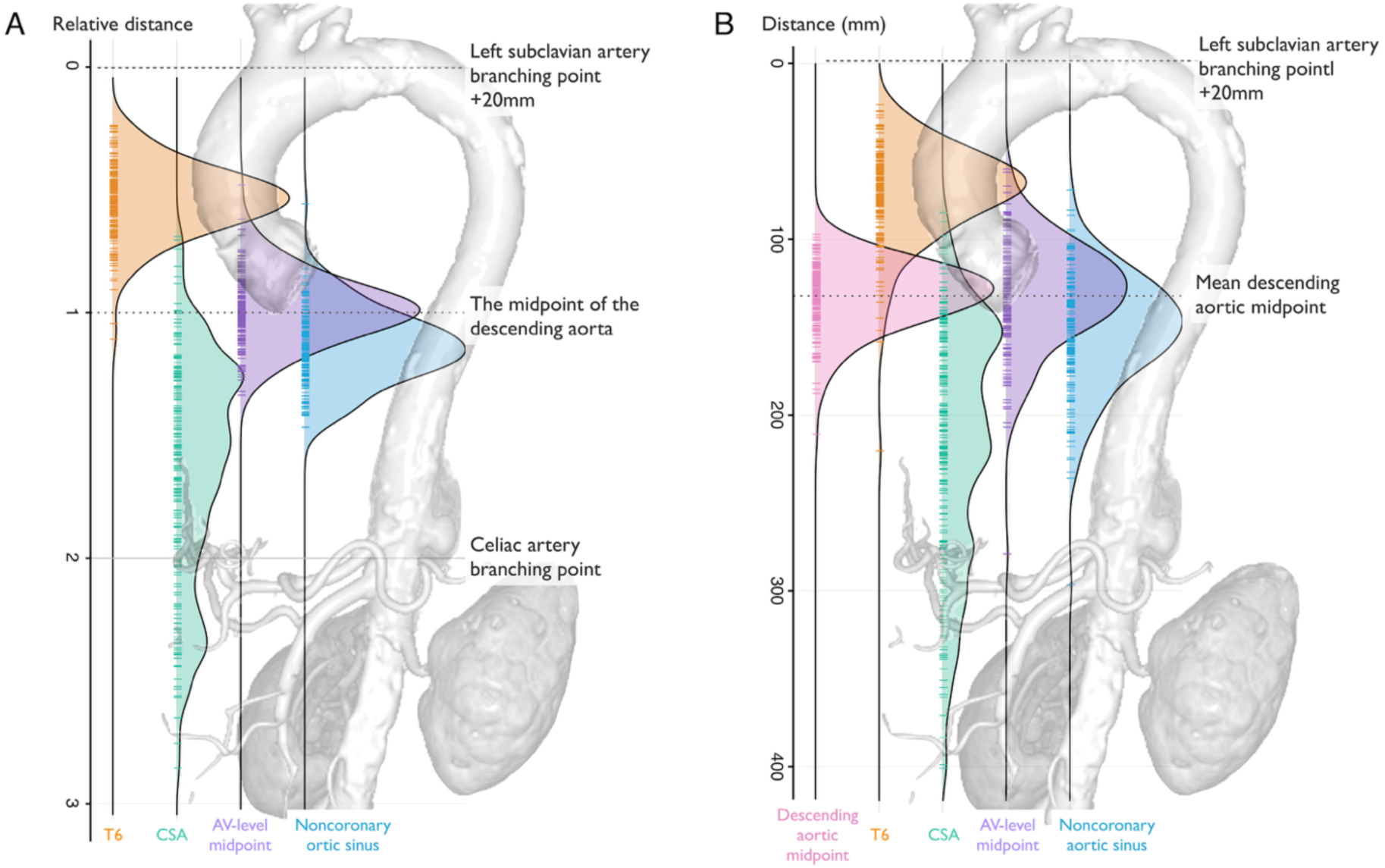
Distribution of candidate landmarks relative to the zone 4 start. Ridgeline plots show candidate-landmark distributions from the zone 4 start, defined as 20 mm distal to the left subclavian artery origin along the aortic centerline. Panel A shows relative distances, with the zone 4 start set to 0, the point immediately proximal to the celiac artery origin set to 2, and the descending aortic midpoint set to 1. Panel B shows absolute distances in millimeters. The AV-level midpoint clustered closest to the descending aortic midpoint, whereas the T6 vertebral plane was more proximal. The CSA origin generally lay distal to the AV-level midpoint, whereas the noncoronary aortic sinus plane overlapped the proximal CSA distribution. AV = aortic valve, CSA = critical segmental artery.

**Table 2.** The distance and relative distance from the zone 4 starting point to each landmark.

| Landmarks | Distance, mm |  |  | Relative distance |  |  |
| --- | --- | --- | --- | --- | --- | --- |
| | Mean ± SD | 95% CI | Median [IQR] | Mean ± SD | 95% CI | Median [IQR] |
| T6 plane | 73.2 ± 26.4 | 69.6 to 76.8 | 70.0 [57.4, 85.8] | 0.54 ± 0.14 | 0.52 to 0.56 | 0.54 [0.46, 0.62] |
| T8 upper border plane | 110.6 ± 27.6 | 106.8 to 114.4 | 106.0 [92.4, 123.3] | 0.83 ± 0.13 | 0.81 to 0.85 | 0.83 [0.75, 0.89] |
| Left coronary aortic sinus plane | 110.3 ± 28.9 | 106.3 to 114.3 | 108.0 [92.5, 122.8] | 0.83 ± 0.14 | 0.81 to 0.85 | 0.83 [0.75, 0.91] |
| AV-level midpoint | 131.1 ± 29.5 | 127.0 to 135.1 | 129.4 [111.6, 146.6] | 0.99 ± 0.14 | 0.97 to 1.00 | 0.99 [0.91, 1.06] |
| Descending aortic midpoint | 132.2 ± 18.2 | 129.7 to 134.7 | 130.4 [119.5, 142.0] | 1.00 | — | — |
| T9 plane | 148.0 ± 30.1 | 143.8 to 152.1 | 143.0 [127.5, 162.1] | 1.11 ± 0.13 | 1.10 to 1.13 | 1.11 [1.04, 1.18] |
| Noncoronary aortic sinus plane | 151.8 ± 31.6 | 147.4 to 156.2 | 149.5 [130.5, 168.8] | 1.14 ± 0.15 | 1.12 to 1.16 | 1.15 [1.05, 1.24] |
| CSA branching point | 216.0 ± 69.2 | 206.5 to 225.6 | 207.5 [156.5, 271.6] | 1.63 ± 0.46 | 1.57 to 1.69 | 1.55 [1.26, 1.93] |
| Celiac branching point | 264.5 ± 36.3 | 259.5 to 269.5 | 260.8 [238.9, 284.0] | 2.00 | — | — |

Note.— The zone 4 starting point was defined as the point 20 mm distal to the left subclavian artery origin along the centerline.
SD = standard deviation, CI = confidence interval, IQR = interquartile range, AV = aortic valve, CSA = critical segmental artery

### Primary Endpoint: AV-Level Midpoint vs Descending Aortic Midpoint

The mean AV-level versus descending aortic midpoint difference was -1.2±18.0 mm (90% CI, -3.3 to 0.9 mm), meeting the prespecified ±20-mm equivalence criterion (P < .001; Figure 4 and Table S2). The 95% limits of agreement for individual differences were -36.5 to 34.1 mm; the absolute difference was ≤20 mm in 151 of 204 patients (74.0%; exact 95% CI, 67.4% to 79.9%). By comparison, the T6 plane was 59.0 mm (95% CI, 56.7–61.3 mm) proximal to the descending aortic midpoint. A T6–aortic midpoint offset of at least 50 mm occurred in 151/204 patients (74.0%; exact 95% CI, 67.4% to 79.9%). In multivariable analysis, older age and aneurysm, compared with dissection, were associated with a more distal AV-level midpoint relative to the aortic midpoint, whereas higher body mass index was associated with a more proximal AV-level midpoint (Table S3).

**Figure 4.**
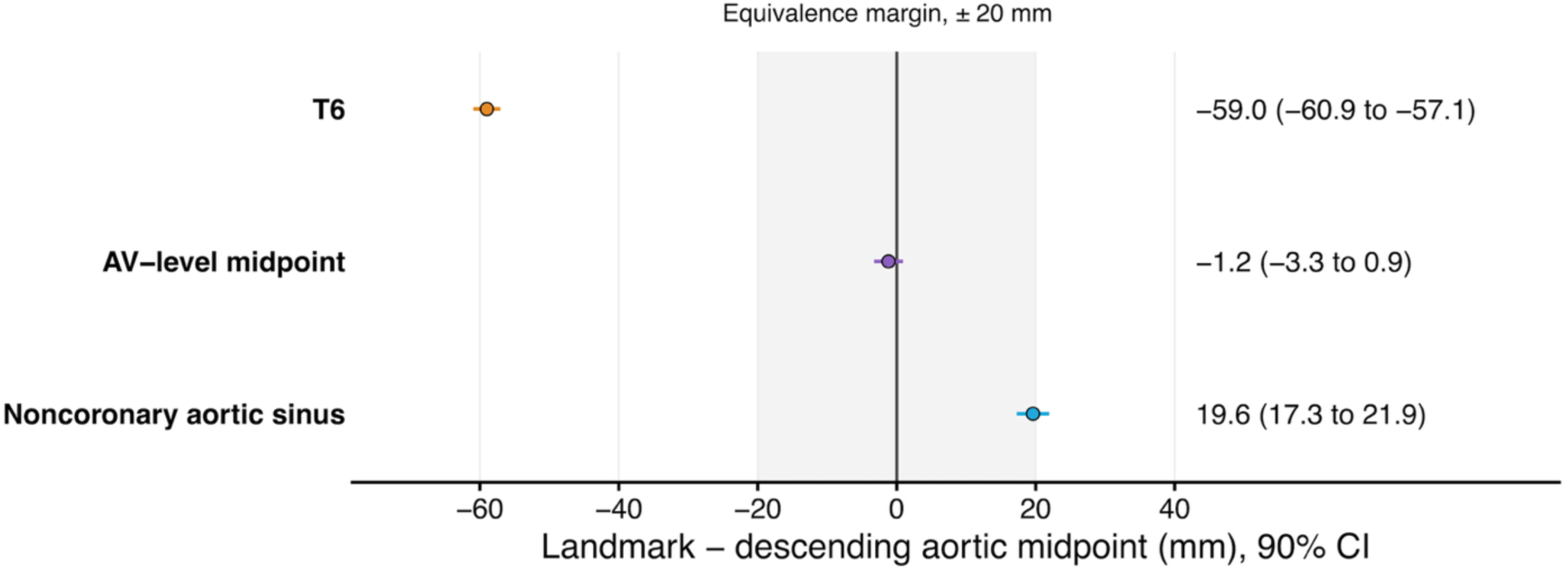
Candidate landmark offsets relative to the descending aortic midpoint. Forest plot shows the mean paired difference between each landmark and the descending aortic midpoint, calculated as landmark distance minus descending aortic midpoint distance, with 90% confidence intervals. The shaded area indicates the prespecified equivalence margin for the AV-level midpoint. Negative values indicate a proximal location relative to the descending aortic midpoint; positive values indicate a distal location. AV = aortic valve.

### Association Between Each Landmark and the CSA Origin

Landmark-to-CSA distances are summarized in Table 3. The CSA origin was distal to the AV-level midpoint in 197 of 204 patients (96.6%; exact 95% CI, 93.1% to 98.6%) and to the descending aortic midpoint in 195 of 204 patients (95.6%; exact 95% CI, 91.8% to 98.0%). Paired classifications did not differ significantly (P = .68) (Figure 5). Distal classification was less frequent for the noncoronary aortic sinus plane (183 patients, 89.7%; exact 95% CI, 84.7% to 93.5%) than for the AV-level midpoint (P = .001) (Table S4). Figure 6 and S2 shows representative cases.

**Figure 5.**
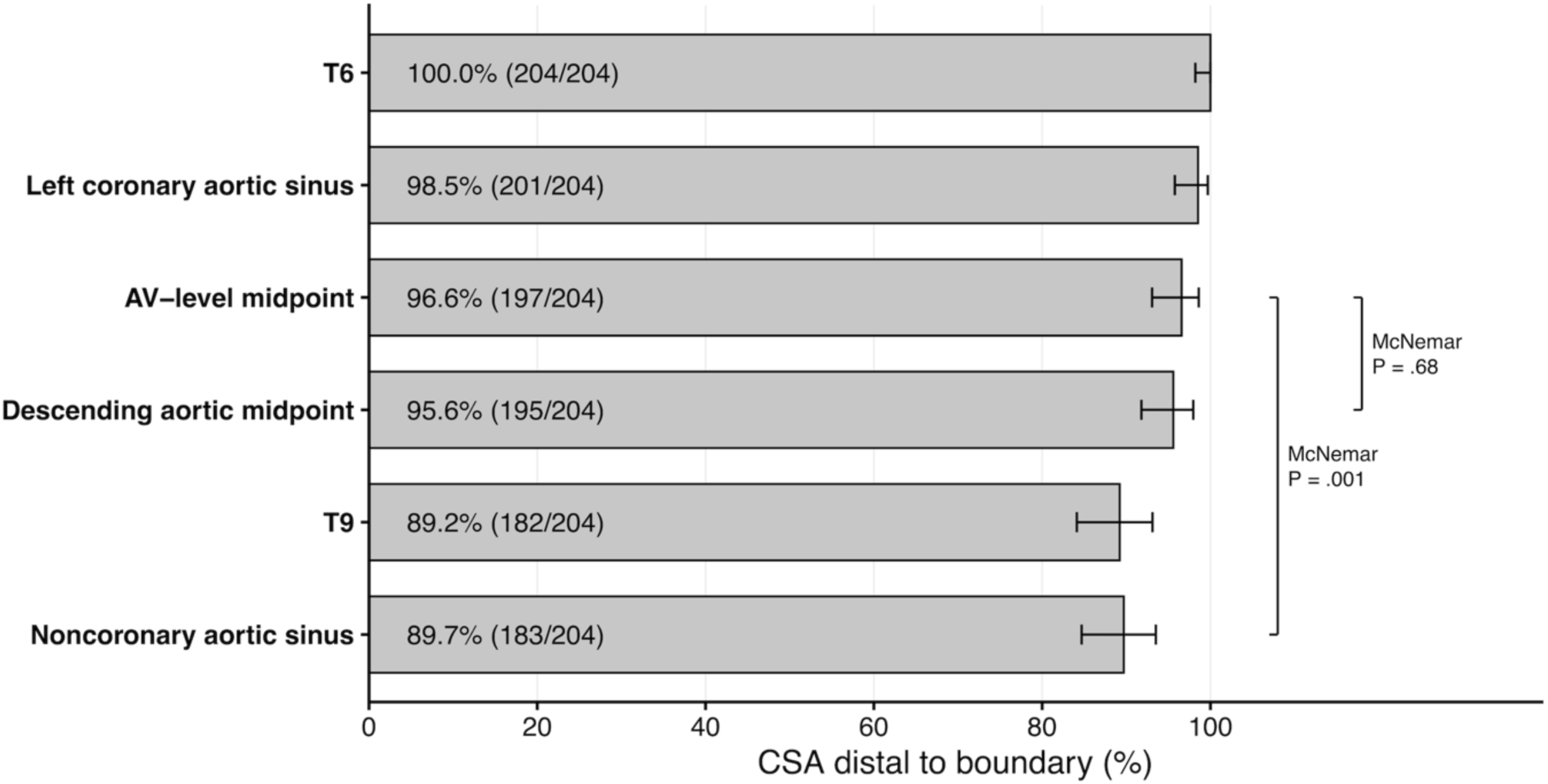
Categorical classification of the CSA origin relative to candidate boundaries. Bars show the proportion of patients with CSA origin distal to each candidate boundary. Error bars indicate exact 95% confidence intervals. Brackets indicate McNemar tests for paired binary classifications. CSA = critical segmental artery.

**Figure 6.**
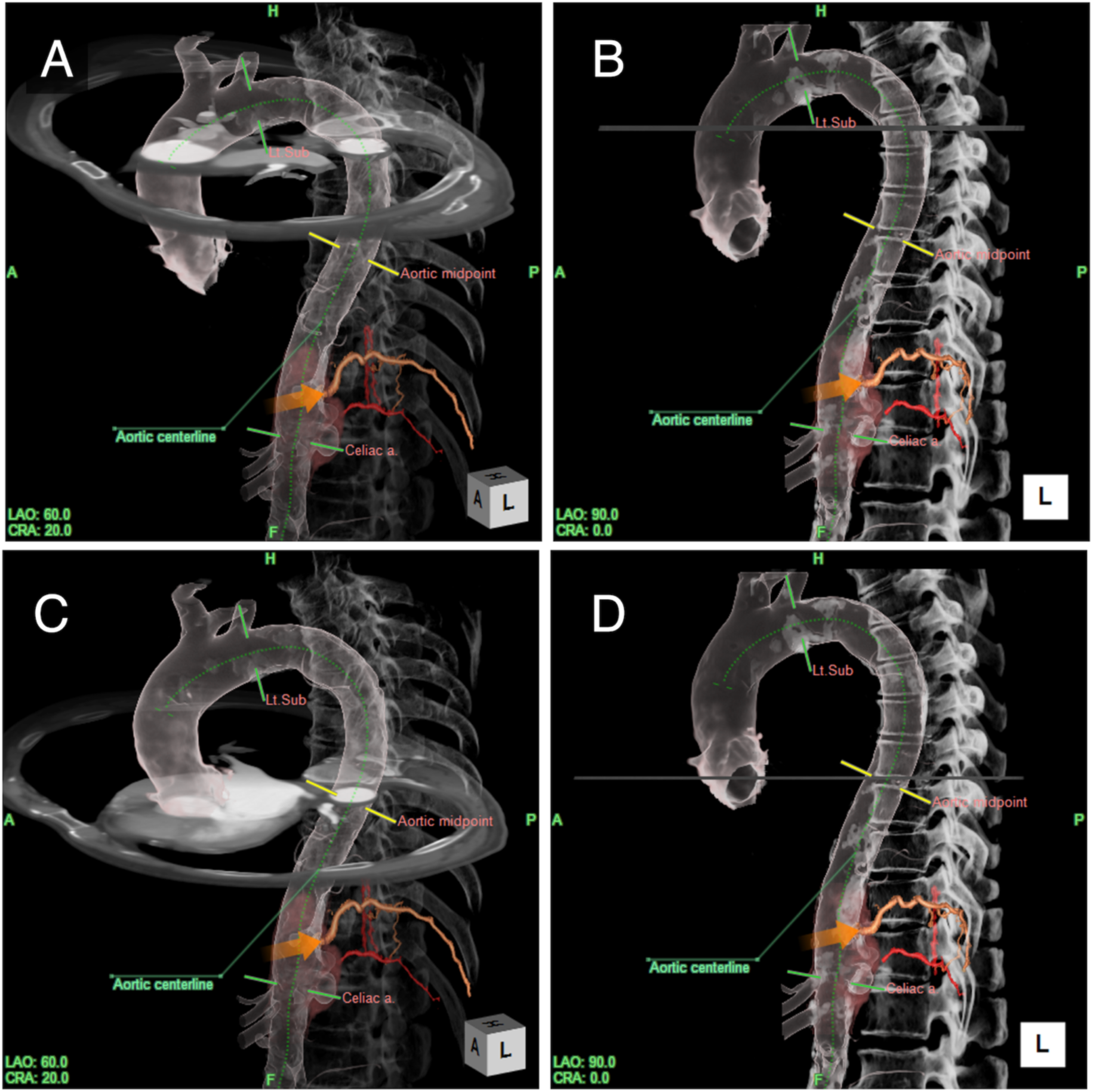
Volume-rendered CT angiography images of a man in his 70s with thoracic aortic aneurysm and penetrating atherosclerotic ulcer are displayed in left anterior oblique (A, C) and left lateral (B, D) views. Gray transverse planes mark evaluated anatomic levels; yellow lines, the descending aortic midpoint; orange arrows, CSA origin; and green dotted lines, the aortic centerline. (A–D) The CSA was the left 11th intercostal artery (orange vessel) and supplied, through an intercostal collateral pathway, the AKA (red vessel), which arose from the left 12th intercostal artery, whose aortic origin was occluded. Panels A and B show the T6 plane, and panels C and D show the AV-level midpoint plane. The T6 and AV- level midpoint planes were 50 mm and 3 mm proximal to the descending aortic midpoint, respectively. AKA = Adamkiewicz artery, AV = aortic valve, CSA = critical segmental artery.

**Table 3.** The distance from each landmark to the CSA branching point.

| Landmarks | Distance, mm |  |  |
| --- | --- | --- | --- |
|  | Mean | 95% CI | Median [IQR] |
| T6 plane | 142.8 | 134.4 to 151.3 | 133.5 [97.2, 193.2] |
| T8 upper border plane | 105.4 | 97.0 to 113.8 | 96.2 [58.6, 152.4] |
| Left coronary aortic sinus plane | 105.7 | 97.5 to 114.0 | 101.8 [56.9, 148.2] |
| AV-level midpoint | 85.0 | 76.7 to 93.2 | 77.4 [37.1, 127.4] |
| Descending aortic midpoint | 83.8 | 75.2 to 92.3 | 72.8 [32.7, 124.0] |
| T9 plane | 68.0 | 59.6 to 76.5 | 60.5 [20.8, 113.8] |
| Noncoronary aortic sinus plane | 64.2 | 55.9 to 72.6 | 54.2 [17.1, 108.1] |

Note.—AV = aortic valve, CSA = critical segmental artery, CI = confidence interval, IQR = interquartile range.

### Exploratory T9 CSA-Origin Analysis

In 51 patients, CSA origin distance correlated with T9 vertebral-level, descending aortic midpoint, and AV-level midpoint distances. The T9 vertebral level had the highest model-fit point estimates, although bootstrap comparisons of standardized beta coefficients showed no significant differences among landmarks (Table S5).

### Measurement Reproducibility

Interobserver and intraobserver intraclass correlation coefficients ranged from 0.94 to 0.99 (Table S6).

## Discussion

Population-level equivalence was observed between the aortic valve-level midpoint and centerline-derived descending aortic midpoint, the zone 4/5 reference (-1.2 mm; 90% confidence interval: -3.3 to 0.9 mm), within the prespecified ±20-mm margin. The T6 vertebral plan was approximately 59 mm proximal, so the descriptions are not anatomically interchangeable. The critical segmental artery origin was distal to the AV-level and descending aortic midpoints in 96.6% (197/204) and 95.6% (195/204), with no significant paired difference (P = .68). The noncoronary aortic sinus plane yielded fewer distal critical segmental artery classifications than the AV-level midpoint (89.7% [183/204]; P = .001), indicating that the AV-level plane requires explicit definition.

The proximity between the AV-level and descending aortic midpoints should be viewed as an empirical population-level relationship rather than a fixed anatomic rule. Several observations support its anatomic plausibility. The descending aorta is constrained by the vertebral column and intercostal arteries(21), whereas the aortic isthmus transitions between the mobile arch and descending aorta(21–23). Cardiac CT studies show the AV/root complex embedded in the cardiac base and coupled to the cardiac fibrous skeleton(24–27). Age-related elongation of the ascending aorta(23,28) and arch(21), with predominant proximal descending aortic involvemen(28), and less distal descending aortic length change(28), provides geometric support. Surface-anatomy and CT studies place the AV/root complex near the mid-sternal body(29); the sternal body spans approximately T4–T5 to T9(30,31), and the descending aorta spans approximately T5 to T12(30), making similar craniocaudal levels plausible. Thus, the AV level and descending aortic midpoint had similar mean population-level positions, but individual differences exceeded ±20 mm in 26.0% of patients, warranting caution in case-by-case boundary determination. In multivariable analysis, older age and aneurysm versus dissection were associated with a more distal AV-level midpoint relative to the descending aortic midpoint, whereas higher body mass index was associated with a more proximal AV-level midpoint. These associations should not be used to localize the offset to either landmark; rather, they may reflect combined effects of age-related aortic elongation(23,28), body habitus-related root/cardiac orientation(24–26), and disease-specific segmental remodeling of the arch and proximal descending aorta(32). Agreement in patients with thoracic aortic disease supports the empirical relevance of the AV level in the intended TEVAR population.

The CSA-origin relationship provides practical context, not an SCI-risk threshold. A prior study found that AKA information changed TEVAR planning or SCI-risk assessment when the distal landing zone extended to zone 5 or beyond, using the mid-descending point as the zone 4/5 boundary(18). Here, the AV-level and descending aortic midpoints had similar CSA distances and distal classifications, whereas classifications varied by vertebral and AV-plane selection. The mid-T9 plane was close to the aortic midpoint by continuous distance but yielded fewer distal CSA classifications (Table S4), underscoring that phrases such as “below T8” require an explicit plane. Because that plane was undefined in the prior repair series(33), the exploratory T8 upper-border result (200/204 [98.0%] distal) may align more closely with a T8-and-lower segmental-territory interpretation than with a T9-plane interpretation. The noncoronary aortic sinus plane was also caudal to the AV-level midpoint and produced fewer distal CSA classifications. The median CSA distance from the zone 4 start was 207.5 mm (IQR, 156.5–271.6), near the 200-mm coverage length used to indicate long-segment higher-risk TEVAR(5,6,34). Thus, coverage extending substantially distal to the AV-level midpoint may provide context for considering AKA imaging(18), and spinal cord protection in patients with other SCI risk factors(7,35).

The AV level may complement vertebral landmarks during TEVAR. Vertebral numbering can be established preoperatively, but repeating it during TEVAR may require table or C-arm movement from the optimal working position; by contrast, the AV level often remains in the procedural field and can be recognized with a catheter, guidewire, or root angiography. In the exploratory T9 analysis, the vertebral level had the highest point estimates, but the descending aortic and AV levels also remained associated with CSA origin distance. These findings suggest complementary roles: vertebral assessment may better support precise segmental localization, whereas the AV level may provide adjunctive procedural orientation. This concept may extend to intraoperative transesophageal echocardiography, where direct vertebral counting is difficult. Because CT images were acquired during deep inspiration with arm elevation, whereas TEVAR is performed under different respiratory and arm-positioning conditions, fluoroscopic AV-level variation may occur. Future fluoroscopic studies should evaluate AV- based landmark identification across respiratory states and patient positions.

This study has limitations. First, this retrospective single-center study included patients who underwent preoperative AKA-CTA, so selection bias and institutional interpretation practices may have influenced the results. Because the cohort included patients with aortic disease and single-time-point imaging, generalizability to normal or milder disease anatomy and longitudinal stability of the AV- aortic midpoint relationship remain uncertain. Second, AKA-CTA was not electrocardiogram-gated, so cardiac motion may have affected the aortic sinus planes; however, high AV-level midpoint measurement reproducibility suggests limited practical effect. Third, the CSA was defined as the largest segmental artery connected to the AKA, without assessment of smaller collateral pathways. Fourth, centerline measurements in dissection followed the true lumen, reflecting the clinically relevant flow lumen and potential device path but differing from the geometric centerline of the entire dissected aorta. Finally, SCI outcomes were not evaluated; therefore, these findings should be interpreted as anatomic validation of a candidate landmark, not validation of an SCI-risk threshold.

In conclusion, the mean aortic valve-level versus descending aortic midpoint difference met the prespecified ±20-mm equivalence criterion in patients with aortic disease, whereas the sixth thoracic vertebral definition showed a systematic proximal shift. Similar relationships of the aortic valve-level and descending aortic midpoints to the critical segmental artery origin supported the anatomic consistency of the aortic valve-level midpoint, while the noncoronary aortic sinus plane findings emphasized the need to define the aortic valve-level plane explicitly.

## List of abbreviations

AV: aortic valve;
AKA: Adamkiewicz artery;
AKA-CTA: Adamkiewicz artery-specific CT angiography;
CSA: critical segmental artery;
TEVAR: thoracic endovascular aortic repair;
SCI: spinal cord ischemia;
CI: confidence interval;
IQR: interquartile range.

## Data Availability

The data used in this study cannot be shared publicly due to privacy concerns for the individuals who participated. The data will be shared upon a reasonable request to the corresponding author.

## Acknowledgments

The authors thank Editage (https://www.editage.jp/) for English language editing. During manuscript preparation and analysis coding (June-July 2026), the authors used ChatGPT (OpenAI, GPT-5.5 and GPT 5.6) and Claude (Anthropic, Fable 5) for language editing, readability improvement, and assistance with R code development and debugging. All AI-assisted outputs, including code suggestions, were reviewed, tested, revised, and edited by the corresponding author. The authors take full responsibility for the final manuscript, analyses, and conclusions. The ideas and interpretations are solely those of the authors.

## Funding information

No funding was received to conduct this study.

## Notes

### Competing Interest Statement

The authors have declared no competing interest.

### Author Declarations

The institutional review board of National Cerebral and Cardiovascular Center approved the study (approval No. R19039-4) and waived written informed consent because of its retrospective design.

