## Supplementary materials for "Aortic Valve Level as a Candidate Zone 4/5 Boundary Landmark in Endovascular Aortic Repair"

**Supplementary file**

**Appendix S1. Details of Anatomic and Derived Definitions**

Aortic centerlines were semiautomatically generated from the aortic arch to the abdominal aorta and manually corrected to follow the aortic lumen. For patients with dissection, centerlines followed the true lumen. Centerline distance increased from proximal to distal. For reported distances, the zone 4 start, defined as 20 mm distal to the left subclavian artery origin along the centerline, was assigned 0 mm.

Anatomic landmarks were classified as branch landmarks or CT axial-plane landmarks. Branch landmarks included the origins of the left subclavian artery, critical segmental artery (CSA), and celiac artery. The CSA origin was defined as the aortic origin of the segmental artery directly giving rise to the Adamkiewicz artery (AKA) or, when that artery was occluded, a segmental artery connected to the AKA through collateral pathways. When multiple candidate segmental arteries were identified, the largest was selected. For each branch landmark, multiplanar reformations orthogonal to the aortic centerline identified the branching plane, and the corresponding centerline distance was recorded.

Axial-plane landmarks included the mid-T6 and mid-T9 vertebral body planes, left coronary aortic sinus plane, and noncoronary aortic sinus plane. Vertebral levels were determined by identifying T12 from rib anatomy and the lumbosacral transition and then counting cranially. The mid-T6 and mid-T9 planes were assigned at the craniocaudal midpoint of the corresponding vertebral body. The left coronary sinus and noncoronary sinus planes were defined as the axial levels of the upper margin of the left coronary aortic sinus and lower margin of the noncoronary aortic sinus of Valsalva, respectively. When either aortic sinus plane fell outside the 160-mm reconstruction field of view, 400-mm field-of-view images were reviewed to confirm axial position. Each axial-plane landmark was converted to centerline distance at the point where its axial z-coordinate intersected the aortic centerline (Figure S1).

The descending aorta was defined by guideline zone 4 and 5 limits, extending from the zone 4 start to immediately proximal to the celiac artery origin. The descending aortic midpoint was the segment centerline midpoint and served as the reference zone 4/5 boundary. The aortic valve (AV)-level midpoint was the midpoint between the centerline distances of the left coronary and noncoronary aortic sinus planes. The descending aortic midpoint, AV-level midpoint, and T6 plane were candidate zone 4/5 boundaries, whereas the two aortic sinus planes were secondary AV-level references.

For exploratory analysis, the T8 upper-border level was estimated as the midpoint between the mid-T6 and mid-T9 centerline distances; this was a derived estimate rather than a directly measured vertebral plane. Relative landmark distances were calculated by normalizing the zone 4 start-to-aortic midpoint distance as 1. Thus, the zone 4 start, aortic midpoint, and celiac artery origin had relative distances of 0, 1, and 2, respectively.

For each landmark, CSA-to-landmark distance was calculated as CSA-origin distance minus landmark distance. Positive values indicated a distal CSA origin, negative values indicated a proximal CSA origin, and 0 indicated the same centerline level. Distal CSA classification was assigned when this difference was greater than 0.

**Supplementary Tables**

**Table S1. Detailed parameters for Adamkiewicz artery-specific CT angiography**

| Parameters | |
|---|---|
| Scanner | SOMATOM Force (Siemens) |
| Collimation, mm | 2 × 128 × 0.6 |
| Scan mode | Dual-power helical scan |
| Tube potential, kV | 70-100 |
| Rotation time, s | 0.5 |
| Pitch | 0.6 |
| Auto exposure control setting | CARE Dose 4D, quality reference mAs 800 at 80 kV for noncontrast scan and 1600 at 80 kV for early- and late-arterial scans |
| Slice thickness, mm | 0.75 |
| Slice interval, mm | 0.5 |
| Field of view, mm | 160 |
| Reconstruction kernel | Bv59 |
| Iterative reconstruction setting | ADMIRE 4 |

Note: The protocol includes noncontrast, early arterial, and late arterial phase acquisitions from the aortic arch to L4. Contrast administration is body-weight-adjusted, with an iodine delivery rate of 26 mg I/kg/s for 18 s, followed by a saline flush. Scan timing is optimized to enhance the T10 level of the descending aorta using the timing-bolus method. Bone-subtraction images are generated using Ziostation2 (Ziosoft).

**Table S2. Distance difference between each landmark and the descending aortic midpoint.**

| Landmarks | Distance, mm | |
|---|---|---|
| | Mean | 90% confidence interval |
| *Vertebral landmarks* | | |
| T6 plane | -59.0 | -60.9 to -57.1 |
| T8 upper border plane | -21.6 | -23.5 to -19.7 |
| T9 plane | 15.8 | 13.7 to 17.9 |
| *Aortic valve landmarks* | | |
| Left coronary aortic sinus plane | -21.9 | -24.0 to -19.8 |
| AV-level midpoint | -1.2 | -3.3 to 0.9 |
| Noncoronary aortic sinus plane | 19.6 | 17.3 to 21.9 |

Note: Values are the mean paired differences between each landmark and the descending aortic midpoint, calculated as landmark minus descending aortic midpoint, with 90% confidence intervals. Negative values indicate a position proximal to the descending aortic midpoint, whereas positive values indicate a distal position.

AV = aortic valve.

**Table S3. Clinical factors associated with the distance difference between the AV-level midpoint and the descending aortic midpoint**

| Predictor | Reference | Standardized beta (95% CI) | P value |
|---|---|---|---|
| Age | | 0.38 (0.23 to 0.53) | <.001 |
| Male sex | Female | -0.04 (-0.21 to 0.12) | .61 |
| Height | | 0.09 (-0.09 to 0.27) | .30 |
| Body mass index | | -0.24 (-0.37 to -0.12) | <.001 |
| Aneurysm | Dissection | 0.22 (0.09 to 0.35) | <.001 |
| Intramural hematoma | Dissection | 0.09 (-0.03 to 0.21) | .15 |
| Other disease | Dissection | 0.06 (-0.06 to 0.19) | .32 |
| Proximal repair | No proximal repair | -0.01 (-0.13 to 0.11) | .87 |

Note: Outcome: AV-level midpoint – descending aortic midpoint, mm; Adjusted $R^2$ for the model = 0.31. AV = aortic valve.

**Table S4. Paired distal CSA classifications for each candidate landmark against the descending aortic midpoint or AV-level midpoint reference**

| Candidate landmark | Cand. only | Ref. only | Both distal | Neither distal | Difference, pp | P value |
|---|---|---|---|---|---|---|
| *Comparison with descending aortic midpoint* | | | | | | |
| AV-level midpoint | 4 | 2 | 193 | 5 | 0.98 | .68 |
| Left coronary aortic sinus plane | 6 | 0 | 195 | 3 | 2.94 | .12 |
| Noncoronary aortic sinus plane | 1 | 13 | 182 | 8 | -5.88 | .01 |
| T8 upper border plane | 6 | 1 | 194 | 3 | 2.45 | .26 |
| T9 plane | 0 | 13 | 182 | 9 | -6.37 | .004 |
| *Comparison with AV-level midpoint* | | | | | | |
| Left coronary aortic sinus plane | 4 | 0 | 197 | 3 | 1.96 | .26 |
| Noncoronary aortic sinus plane | 0 | 14 | 183 | 7 | -6.86 | .001 |
| T8 upper border plane | 4 | 1 | 196 | 3 | 1.47 | .37 |
| T9 plane | 0 | 15 | 182 | 7 | -7.35 | .001 |

Note: Each entry indicates the number of patients classified by whether the CSA origin was distal to the candidate landmark and/or the reference landmark within the same patient. Cand. only indicates distal classification by the candidate landmark but not by the reference landmark; Ref. only indicates distal classification by the reference landmark but not by the candidate landmark. Difference is expressed in percentage points and calculated as the candidate-landmark distal percentage minus the reference-landmark distal percentage. P values are from McNemar tests for paired binary classifications with Holm adjustment. AV = aortic valve; CSA = critical segmental artery.

**Table S5. Results of the exploratory T9 CSA-origin analysis**

| Candidate landmark | Pearson r | Adjusted standardized beta | Model P value | Standardized beta difference vs T9 | Difference P value |
|---|---|---|---|---|---|
| T9 plane | 0.93 (0.87 to 0.96) | 0.91 (0.78 to 1.04) | <.001 | - | - |
| Descending Aortic midpoint | 0.89 (0.81 to 0.93) | 0.88 (0.73 to 1.03) | <.001 | -0.03 (-0.16 to 0.08) | .51 |
| AV-level midpoint | 0.76 (0.62 to 0.86) | 0.86 (0.67 to 1.05) | <.001 | -0.07 (-0.23 to 0.11) | .44 |

Note: Pearson r and standardized beta values are shown with 95% confidence intervals. The outcome was CSA origin distance from the zone 4 starting point. Candidate landmark distances were evaluated in separate models to avoid overinterpreting collinear landmarks. Adjusted models included CSA side, age, sex, height, body mass index, dissection versus non-dissection, and proximal treatment. Standardized beta coefficients were calculated for each candidate landmark term. Standardized beta differences are calculated as the candidate-landmark standardized beta minus the T9 vertebral-plane standardized beta. Bootstrap comparisons of standardized beta differences used 10,000 row-level resamples.

AV = aortic valve; CSA = critical segmental artery.

**Table S6. The inter- and intra-observer agreements**

| Landmarks | Inter-observer | Intra-observer |
|---|---|---|
| Descending Aortic midpoint | 0.960 (0.843 to 0.986) | 0.994 (0.988 to 0.998) |
| AV-level midpoint | 0.944 (0.766 to 0.981) | 0.989 (0.976 to 0.995) |
| T6 plane | 0.950 (0.890 to 0.978) | 0.991 (0.981 to 0.996) |
| CSA branching point | 0.993 (0.980 to 0.997) | 0.997 (0.994 to 0.999) |

Note: Intraclass correlation coefficients and 95% confidence intervals are shown.
AV = aortic valve; CSA = critical segmental artery.

**Supplementary Figures**

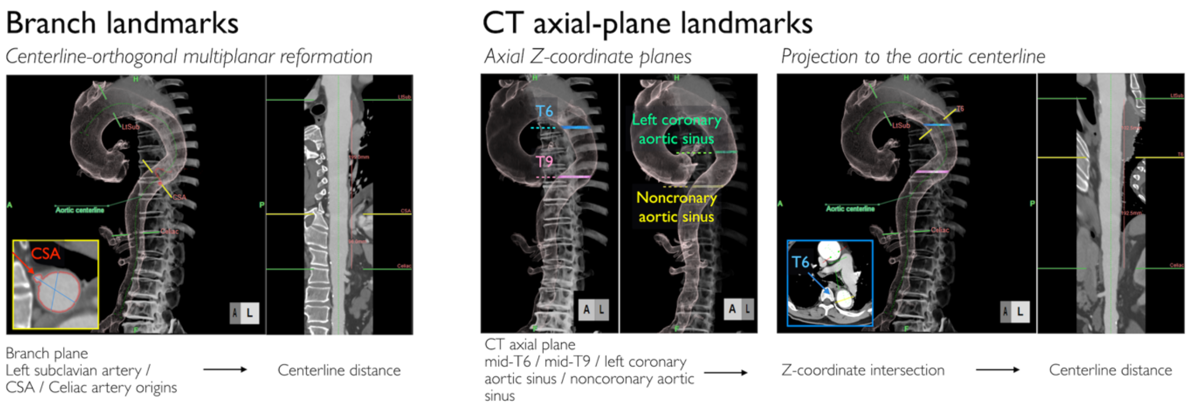


**Figure S1. The conversion of anatomical landmarks to aortic centerline distances.**
Branch landmarks were identified on multiplanar reformations orthogonal to the aortic centerline, and the corresponding branch plane was recorded as a centerline distance. CT axial-plane landmarks, including the mid-T6 and mid-T9 vertebral body planes and the left coronary and noncoronary aortic sinus planes, were assigned to the centerline distance at which the axial z-coordinate intersected the aortic centerline.
AV = aortic valve.

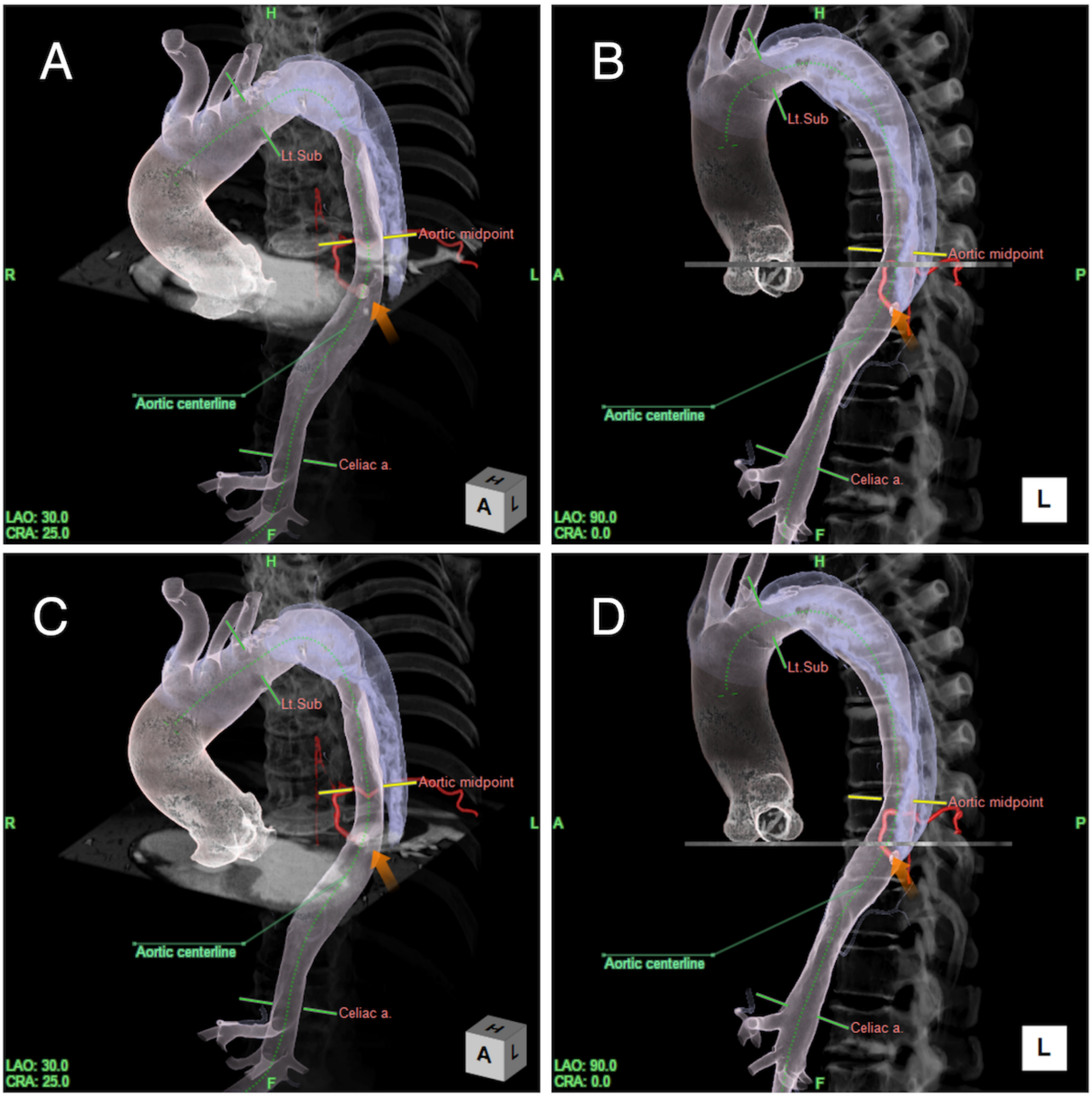


**Figure S2. Volume-rendered CT angiographic images of a woman in her 80s with aortic dissection are shown in left anterior oblique (A, C) and left lateral (B, D) views.**

Gray transverse planes mark evaluated anatomic levels; yellow lines, the descending aortic midpoint; orange arrows, CSA origin; and green dotted lines, the aortic centerline. The CSA was the left ninth intercostal artery (red vessel). Panels A and B show the AV-level midpoint plane, and panels C and D show the noncoronary aortic sinus plane. The AV-level midpoint and noncoronary sinus planes were 5 mm and 25 mm distal to the descending aortic midpoint, respectively. The CSA origin was 23 mm distal to the AV-level midpoint but only 3 mm distal to the noncoronary aortic sinus plane.
